# Associations between structural brain changes and blood neurofilament light chain protein in treatment-resistant schizophrenia

**DOI:** 10.1101/2024.04.07.24305362

**Authors:** Brandon-Joe Cilia, Dhamidhu Eratne, Cassandra Wannan, Charles Malpas, Shorena Janelidze, Oskar Hansson, Ian Everall, Chad Bousman, Naveen Thomas, Alexander F Santillo, Dennis Velakoulis, Christos Pantelis

**Author notes:** **Corresponding Author** Brandon-Joe Cilia Neuropsychiatry, The Royal Melbourne Hospital, 300 Grattan Street, Parkville, Melbourne VIC 3050, Australia.

## Abstract

**Background and Hypothesis:** Around 30% of people with schizophrenia are refractory to antipsychotic treatment (treatment-resistant schizophrenia; TRS). While abnormal structural neuroimaging findings, in particular volume and thickness reductions, are often observed in schizophrenia, it is anticipated that biomarkers of neuronal injury like neurofilament light chain protein (NfL) can improve our understanding of the pathological basis underlying schizophrenia. The current study aimed to determine whether people with TRS demonstrate different associations between plasma NfL levels and regional cortical thickness reductions compared with controls.

**Study Design:** Measurements of plasma NfL and cortical thickness were obtained from 39 individuals with TRS, and 43 healthy controls. T1-weighted magnetic resonance imaging sequences were obtained and processed via FreeSurfer. General linear mixed models adjusting for age and weight were estimated to determine whether the interaction between diagnostic group and plasma NfL level predicted lower cortical thickness across frontotemporal structures and the insula.

**Study Results.:** Significant (false discovery rate corrected) cortical thinning of the left (*p* = 0.001, _η_*^2^_p_* = 0.104) and right (*p* < 0.001, _η_*^2^* = 0.167) insula was associated with higher levels of plasma NfL in TRS, but not in healthy controls.

**Conclusions.:** The association between regional thickness reduction of the insula bilaterally and plasma NfL may reflect a neurodegenerative process during the course of TRS. The findings of the present study suggest that some level of cortical degeneration localised to the bilateral insula may exist in people with TRS, which is not observed in the normal population.

## Introduction

Schizophrenia is a complex and lifelong neuropsychiatric illness affecting nearly 1% of people worldwide. The most enduring model posits that schizophrenia is a neurodevelopmental disorder and postulates that susceptibility genes in combination with environmental risk factors occurring in early life produce a neurodevelopmental lesion that results in schizophrenia two to three decades later.(1–4) However, recent longitudinal neuroimaging studies showing progressive brain changes over the course of the illness, have revived the concept of schizophrenia as a neuroprogressive, perhaps even neurodegenerative, brain disorder in at least a subset of patients with severe disorder.(5,6)

Up to one in three people with schizophrenia inadequately respond to conventional antipsychotic treatment and are described as having treatment-resistant schizophrenia (TRS).(7) TRS is broadly defined as the absence of symptomatic improvement despite adherence to ≥2 different antipsychotic trials of appropriate dosage and duration.(8) People with TRS also tend to have significant residual symptoms, a worse prognosis, and a greater level of social and occupational dysfunction compared to those who respond better to antipsychotic treatment.(9)

Neuroimaging studies implicate widespread brain structural abnormalities in schizophrenia throughout the course of the disease, including in chronic schizophrenia, at first-episode psychosis, and even among cohorts at high risk for psychosis.(10–15) Among the most widely replicated neuroimaging findings in schizophrenia is cortical volume loss, in particular, of frontal and temporal areas.(16,17) Cortical volume, which is by definition the product of cortical thickness and cortical surface area, may erroneously confound these two separate and partly unrelated neuroanatomical features. Emerging evidence suggests that although cortical thickness and surface area are both heritable features, they are likely under distinct and independent sets of genetic and phenotypic influences.(18,19) Surface area is understood to be predominantly under neurodevelopmental and genetic influence whereas cortical thickness tends to be more affected by neurodegenerative and environmental factors.(15,20) Regional and global cortical thickness reductions have also been demonstrated in schizophrenia, implicating cortical thinning in the neuropathology and symptomatology of the illness.(12,15,21–23) It has therefore been postulated that findings of abnormal brain volumes in schizophrenia may be predominantly driven by cortical thickness changes rather than surface area loss.(15) Taken together, changes to cortical thickness may better describe the aetiological and pathological (perhaps neurodegenerative) processes driving disease progression in schizophrenia.

As TRS is often associated with structural neuroimaging abnormalities,(13,34–36) it is plausible that cortical volume and thickness reductions may be associated with elevations of markers of neuroaxonal injury. A novel and minimally invasive approach to assessing neuronal integrity involves measurement of fluid biomarkers. Neurofilaments are a family of neuron-specific cytoplasmic proteins that confer stability to neuronal axons.(24,25) Neurofilament light chain protein (NfL) is the lowest molecular weight, most soluble, and most abundant of the neurofilament protein family.(26) Large-calibre myelinated axons are particularly enriched in NfL, and as a marker of neuronal injury/degeneration, numerous studies have demonstrated elevated cerebrospinal fluid (CSF) and blood levels of NfL in a broad range of neurological and neurodegenerative disorders.(24,26–29) Several studies to date have investigated NfL in schizophrenia.(30–33) Despite largely negative findings at the group level, disease-driven axonal injury may instead be a feature of a subset of people with psychoses, such as those with more severe phenotypes.

The primary aim of this study was to investigate the association between plasma NfL levels and cortical thickness in people with TRS compared to matched controls. Brain regions of interest included frontal, temporal, and the insula. The primary hypothesis was that raised plasma NfL levels will be associated with cortical thinning of these brain structures in TRS patients, but not among healthy controls.

## Methods and materials

### Participants

Samples and data were made available to the current study on behalf of the Cooperative Research Centre (CRC) for Mental Health. Data collection for the psychosis cohort including clinical assessment information, blood sampling and neuroimaging was conducted by the Melbourne Neuropsychiatry Centre (Department of Psychiatry, University of Melbourne). We have reported NfL values for this cohort previously.(33) The CRC Psychosis Study protocol and this project were approved by the Melbourne Health Human Research Ethics Committee (ethics IDs 2012.069 and 2020.142, respectively). All participants provided informed, written consent before participating.

Individuals with TRS were recruited from a range of inpatient and outpatient psychiatric services across Melbourne, Australia from 2012 to 2017 as part of the CRC Psychosis Study. Treatment-resistance was defined by current clozapine use and inclusion criteria for the TRS group included a schizophrenia diagnosis and being aged 18 to 65 years. A comparison group of unrelated, age- and sex-matched controls who did not have schizophrenia was also drawn from the same catchment site as the patient group.

All recruited patients underwent a process of comprehensive psychopathology and neuropsychology assessments as well as a medical history. The MINI International Neuropsychiatric Interview(37) was administered to all participants in order to confirm a diagnosis of schizophrenia in the patient group and to rule out psychiatric disorders in healthy controls. Additional clinical measures were recorded for both groups including other medical comorbidities, a family history of mental illness, and substance abuse history.

### Neurofilament light chain protein assay protocol

Fasting blood samples were obtained from all participants and plasma was stored at −80°C. Plasma concentration of NfL was measured using a single molecule array NF-Light Advantage Kit (SR-X) according to the manufacturer’s instructions (Quanterix Corporation, Lexington, MA USA). SR-X digital immunoassays have a mean limit of detection of 0.0552 pg mL^−1^. All samples were diluted 1:4. Four internal control samples of pooled plasma were included in every plate. The mean intra-plate coefficient of variability (CV) was 4.97%, and the average inter-plate CV was 6.59%.

Two extreme outliers were noted, one in the control group (45-year-old male, plasma NfL 46.2 pg mL^−1^, z-score = 6.28) and another in the TRS group (48-year-old female, plasma NfL 21.4 pg mL^−1^, z-score = 3.98). A generally accepted cut-off value for extreme values is a z-score of 3 or more. As both outliers were more than three standard deviations above the means of their respective groups, they were excluded from all analyses. Outlier samples were measured three times, and levels did not differ by more than 10% each time.

### Image data acquisition

Magnetic resonance imaging (MRI) sequences were acquired in a Siemens Avanto 3T Magnetom TIM Trio scanner. For each participant, a T1-weighted Magnetisation-Prepared Rapid Acquisition Gradient Echo sequence was obtained with the following sagittal imaging parameters: 176 sagittal slices of 1 mm thickness without gap, field of view of 250 x 250 mm^2^, repetition time of 1980 ms, echo time of 4.3 ms, flip angle of 15°, and an acquisition matrix of 256 x 256. A final reconstructed voxel resolution of 0.98 x 0.98 x 1.0 mm^3^ was achieved.

### Image processing and estimation of cortical thickness

MRI data was processed with the FreeSurfer software package, version 6.0 (https://surfer.nmr.mgh.harvard.edu/) to estimate cortical thickness. FreeSurfer automatically labels the surface of the brain to construct a cortical map based on probabilistic information.(38,39) In brief, the automated processing pipeline consisted of intensity normalisation, skull stripping, automated Talairach transformation, and corrections for motion, signal intensity and topology.(39–44) Next, cortical surfaces were reconstructed. This was achieved by overlaying grey-white matter boundaries to form the white matter surface layer, and grey-CSF boundaries to create the pial surface layer—the cortex being the region defined by the white matter and pial boundaries.(40) All reconstructed surfaces were visually checked for defects in skull stripping, intensity normalisation, white-grey matter segmentation and motion artefact by a qualified technician. Errors in boundary demarcation were manually corrected via FreeSurfer’s native editing tools, following a standardised editing protocol. Edited images were reprocessed, and the output visually inspected for inaccuracies again. These steps were repeated until all surface errors were corrected. The cortical surface was then parcellated into 68 regions according to the Desikan-Killiany atlas.(45) An estimate of the average cortical thickness for each region was obtained by averaging across all vertices of the given region for each subject.

### Statistical analysis

All statistical analyses were performed using the Statistical Package for the Social Sciences (SPSS) versions 28 & 29.

#### Analysis of demographic measures

Group differences of demographic (such as age at sample, sex, and years of schooling) and clinical (such as illness duration, and degree of functional impairment) measures were compared using chi-square tests for categorical variables, and independent samples t-tests for continuous variables. Statistical significance was defined as *p* < 0.05. Missing data was dealt with using pairwise deletion.

#### Analysis of between-group differences in plasma neurofilament light chain protein level and cortical thickness

Independent samples t-tests were used to compare levels of plasma NfL between the healthy controls and people with TRS.

Group differences in cortical thickness were examined using ANCOVAs, with age and weight used as covariates. Results were then corrected for multiple comparisons in each hemisphere using a false discovery rate (FDR) of *p* < 0.05.(47)

#### Analysis of interactions between plasma neurofilament light chain protein level and group

To determine whether differential relationships exist between plasma NfL levels and cortical thickness by group, a series of general linear mixed models (GLMMs) with random effect of clinical site were estimated. GLMMs in the present study were all adjusted for age given its relationship with plasma NfL levels.(24) Weight was also included as a covariate due to emerging evidence of a negative association between plasma NfL levels and weight which has been attributed to the dilutional effect of increased blood volume.(46) Each model included the main effects of group membership and plasma NfL level, in addition to age and weight as covariates. The interaction term (NfL-by-Group) was included in the model to investigate whether NfL- and group-related differences of the thickness measures were significant. In brief, NfL-by-Group determines whether membership in either diagnostic group (i.e., healthy control or TRS) significantly modifies the association between plasma NfL and thickness of each brain structure.

The brain structures that were investigated in the present study accounted for all frontotemporal regions as well as the insula as these cortical structures are most consistently implicated in schizophrenia neuroimaging studies.(15,22,34) The ENIGMA Schizophrenia Working Group identified a loco-regional pattern of cortical thinning across 4474 patients with schizophrenia compared to 5098 healthy controls, with the largest effect sizes in frontal and temporal areas.(15) Zhao et al. (2022) similarly demonstrated cortical thickness reductions across the frontotemporal areas, as well as the right insula in a cohort of people with chronic schizophrenia.(22) A complete list of cortical regions included in the present study is available in Table S1 in the supplement.

Correction for multiple comparisons was made using a FDR of *p* < 0.05.(47) Hence, significance for the NfL-by-Group interaction would indicate that the trendlines between cortical thickness and plasma NfL level are significantly different between the two groups, when adjusted for age and weight. Bias-corrected and accelerated (BCa) confidence intervals (CI) were generated for all GLMMs via bootstrapping using 2000 replicates. Partial eta squared (_η_*^2^_p_*) was also reported to give a measure of effect size for the NfL-by-Group interaction term and the main effects of plasma NfL.

## Results

### Sample characteristics

A total of 82 participants from the initial pool of volunteers recruited to the CRC Psychosis Study Treatment Resistant Schizophrenia Biobank had both plasma samples as well as MRI scans available. Full demographic and clinical variables are presented in Table 1. Thirty-nine participants had a diagnosis of TRS and a group of 43 control participants were also included in the study. The TRS group had a mean illness duration of 16.3 years with an average illness onset at 21.6 years. The TRS and control groups did not differ in age (*p* = 0.433) nor in sex (*p* = 0.097).

**Table 1.** Demographic and clinical characteristics.

|  | Control | TRS |
| --- | --- | --- |
| *N* | 43 | 39 |
| Age at sample, years | 39.6 [36.1, 43.1] | 37.9 [35.1, 40.6] |
| Sex, *n* female (%) | 16 (37.2%) | 8 (20.5%) |
| Age at onset, years | – | 21.6 [19.6, 23.6] (n=38) |
| Duration of illness, years | – | 16.3 [14.2, 18.4] (n=38) |
| PANSS total | 29.7 [29.0, 30.4] | 58.7 [54.3, 63.1] |
| *PANSS positive* | 7.2 [7.0, 7.4] | 15.2 [13.2, 17.2] |
| *PANSS negative* | 6.7 [6.3, 7.1] | 16.3 [14.3, 18.3] |
| *PANSS general* | 15.7 [15.4, 16.1] | 27.2 [25.4, 29.1] |
| Years of schooling | 15.9 [15.0, 16.9] | 12.2 [11.2, 13.3] |
| Current smoker (last 12 months) | 7 (16.2%) | 15/36 (41.7%) |
| Diagnosis of alcohol use disorder | 6 (14.0%) | 9 (23.1%) |
| Other substance use disorder | 4 (9.3%) | 11 (28.2%) |
| Hypertension | 1 (2.3%) | 4/38 (10.5%) |
| Diabetes | 0% | 3/38 (7.9%) |
| Dyslipidaemia | 0% | 5 (12.8%) |
| Weight, kg | 73.5 [70.4, 76.6] | 95.4 [88.7, 102.2] |
| Current IQ | 111.7 [107.2, 116.2] (n=40) | 88.5 [82.3, 94.7] (n=38) |
| Clozapine level, µg/mL | – | 464.9 [379.3, 550.6] (n=38) |
| History of head injury | 7 (16.3%) | 18 (46.2%) |
| Epilepsy | 0% | 7 (17.9%) |
| Other neurological disorders | 0% | 8 (20.5%) |
| Family history of mental illness | 13 (30.2%) | 20 (51.3%) |
| *psychotic* | 3 (7.0%) | 14 (35.9%) |
| *non-psychotic* | 10 (23.3%) | 6 (15.4%) |
| Problems with birth | 4/42 (9.5%) | 10/38 (26.3%) |
| Ever had ECT | 0% | 9/37 (24.3%) |

*Note:* Data reported as mean [95% CI] or *n* (%).
ECT, electroconvulsive therapy; PANSS, Positive and Negative Syndrome Scale; TRS, treatment-resistant schizophrenia.

Individuals with TRS completed fewer years of schooling (*p* < 0.001) and had lower current IQ scores (*p* < 0.001) compared to matched controls. People with TRS were also more likely to have comorbidities like epilepsy (*p* = 0.004) and dyslipidaemia (*p* = 0.015) versus control subjects. Tobacco and other substance use disorders were more common among participants with TRS, although alcohol use was not different between the two groups (*p* = 0.286). People with TRS were also more likely to have had a previous head injury than controls (*p* = 0.003).

Twenty participants with TRS (51.3%) reported having a family history of mental health disorders, with psychotic illnesses being the largest contributor. The opposite trend was observed in healthy controls where non-psychotic disorders (23.3%) predominated among those with a family history of mental illness. 24.3% of TRS participants (9/37) had received electroconvulsive therapy.

### Group differences in neurofilament light chain protein

At the group level, there was no significant mean difference between levels of plasma NfL in people with TRS compared to controls (Table 2). Controls had a mean NfL of 6.1 pg/mL compared to 5.5 pg/mL recorded for people with TRS.

**Table 2.** Between-group comparisons of plasma neurofilament light chain protein level.

|  | Control | TRS |
| --- | --- | --- |
| *N* | 43 | 39 |
| Age at sample, years | 39.6 (11.4) | 37.9 (8.4) |
| Sex, *n* female (%) | 16 (37.2%) | 8 (20.5%) |
| Plasma NfL, pg/mL | 6.1 [5.3, 6.8] | 5.5 [4.6, 6.4] |

*Note:* Data reported as mean [95% CI] or *n* (%). CI, confidence interval; NfL, neurofilament light chain protein.

### Group differences in cortical thickness

In comparison with controls, TRS participants were found to have reduced cortical thickness of the insula bilaterally as well as all frontotemporal structures, except for the transverse temporal bilaterally, right pars triangularis, left pars orbitalis, and left rostral middle frontal regions (Table 3).

**Table 3.** Between-group comparisons of cortical thickness.

| Hemisphere | Structure | Control |  | TRS |  | Control vs. TRS |  |
| --- | --- | --- | --- | --- | --- | --- | --- |
| | | Mean | SD | Mean | SD | *F*-<br>statistic | $\eta^2_p$ |
| Left | Caudal middle frontal | 2.604 | 0.122 | 2.481 | 0.132 | 22.650* | 0.227 |
|  | Lateral orbitofrontal | 2.657 | 0.103 | 2.571 | 0.121 | 12.885* | 0.143 |
|  | Medial orbitofrontal | 2.421 | 0.115 | 2.377 | 0.126 | 7.184* | 0.085 |
|  | Entorhinal | 3.508 | 0.254 | 3.263 | 0.318 | 10.000* | 0.115 |
|  | Pars opercularis | 2.625 | 0.115 | 2.531 | 0.139 | 15.406* | 0.167 |
|  | Pars orbitalis | 2.647 | 0.161 | 2.573 | 0.176 | 3.513 | 0.044 |
|  | Pars triangularis | 2.475 | 0.159 | 2.421 | 0.167 | 7.351 | 0.087 |
|  | Rostral middle frontal | 2.351 | 0.111 | 2.322 | 0.122 | 3.644 | 0.045 |
|  | Superior frontal | 2.742 | 0.124 | 2.607 | 0.120 | 38.999* | 0.336 |
|  | Precentral gyrus | 2.668 | 0.141 | 2.576 | 0.156 | 9.351* | 0.108 |
|  | Inferior temporal | 2.845 | 0.129 | 2.704 | 0.113 | 23.676* | 0.235 |
|  | Middle temporal | 2.915 | 0.115 | 2.792 | 0.119 | 26.293* | 0.255 |
|  | Superior temporal | 2.863 | 0.139 | 2.781 | 0.158 | 7.539* | 0.089 |
|  | Transverse temporal | 2.523 | 0.213 | 2.437 | 0.232 | 3.019 | 0.038 |
|  | Insula | 3.030 | 0.163 | 2.922 | 0.182 | 9.656* | 0.111 |
| Right | Caudal middle frontal | 2.528 | 0.105 | 2.389 | 0.129 | 25.187* | 0.246 |
|  | Lateral orbitofrontal | 2.605 | 0.119 | 2.471 | 0.136 | 25.782* | 0.251 |
|  | Medial orbitofrontal | 2.356 | 0.128 | 2.56 | 0.161 | 9.622* | 0.111 |
|  | Entorhinal | 3.580 | 0.362 | 3.343 | 0.432 | 6.193* | 0.074 |
|  | Pars opercularis | 2.560 | 0.143 | 2.482 | 0.141 | 6.467* | 0.077 |
|  | Pars orbitalis | 2.626 | 0.141 | 2.487 | 0.152 | 18.367* | 0.193 |
|  | Pars triangularis | 2.411 | 0.140 | 2.357 | 0.154 | 3.121 | 0.039 |
|  | Rostral middle frontal | 2.230 | 0.091 | 2.192 | 0.102 | 5.024* | 0.061 |
|  | Superior frontal | 2.659 | 0.118 | 2.512 | 0.113 | 31.918* | 0.293 |
|  | Precentral gyrus | 2.595 | 0.143 | 2.510 | 0.162 | 7.160* | 0.085 |
|  | Inferior temporal | 2.859 | 0.118 | 2.756 | 0.149 | 11.789* | 0.133 |
|  | Middle temporal | 2.903 | 0.116 | 2.810 | 0.126 | 15.556* | 0.0168 |
|  | Superior temporal | 2.855 | 0.142 | 2.782 | 0.170 | 5.169* | 0.063 |
|  | Transverse temporal | 2.462 | 0.215 | 2.425 | 0.241 | 0.705 | 0.009 |
|  | Insula | 3.085 | 0.132 | 2.974 | 0.139 | 14.103* | 0.155 |

*Note:* \* $p < 0.05$ , FDR-corrected.
SD, standard deviation; TRS, treatment-resistant schizophrenia.

### Interactions between neurofilament light chain protein and cortical thickness

Region of interest (ROI) analysis of frontotemporal and insula cortical thicknesses showed a significant NfL-by-Group interaction in the left and right insula (*p* = 0.001, and *p* < 0.001, FDR-corrected, respectively). This was interpreted as a statistically significant difference in the relationship between plasma NfL and cortical thickness of the bilateral insula across the two groups. Specifically, a higher plasma NfL level significantly were associated with lower cortical thickness of these structures among participants with TRS but not in controls. Interaction plots of the left and right insula with regression lines and 95% CIs are presented in Figure 1. Trendlines and CIs are separated by diagnostic group (i.e., control or TRS).

**Figure 1.**
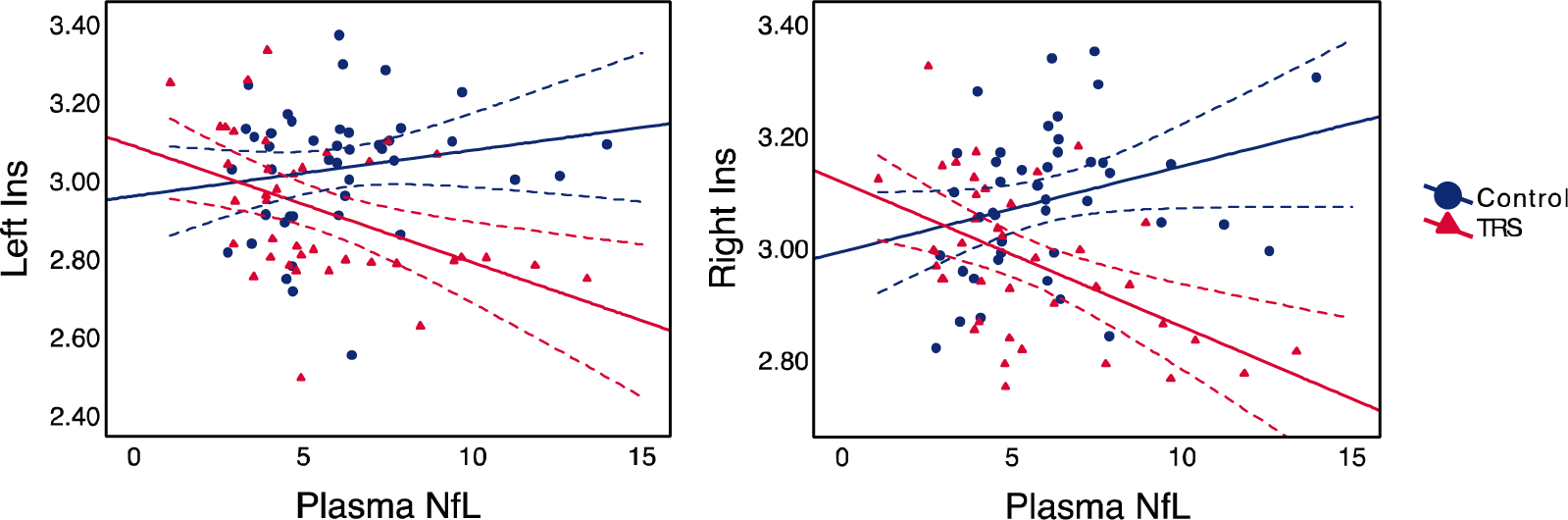
Interaction plots with trendlines and 95% confidence intervals for the left and right insula. Plasma NfL in pg/mL; thickness in mm. *Note:* Ins, insula; NfL, neurofilament light chain protein.

Other biologically plausible regions such as the left middle and inferior temporal gyri, while not significant regions following FDR-correction, however these showed similar effect sizes. Plasma NfL did not predict differences in cortical thickness between people with TRS and controls in other brain structures (Supplementary Figures S1 & S2). Complete results are presented in Table 4.

**Table 4.** General linear mixed models and their corresponding plasma NfL-by-Group interaction term for each brain structure thickness.

| Hemisphere | GLMM | Plasma NfL-by-Group interaction |  |  |  |
| --- | --- | --- | --- | --- | --- |
| | | B* | BCa 95% CI | $\eta^2_p$ | p-value |
| Left | Caudal middle frontal | 0.010 | [-0.009, 0.027] | 0.011 | 0.265 |
|  | Lateral orbitofrontal | 0.014 | [-0.003, 0.035] | 0.029 | 0.100 |
|  | Medial orbitofrontal | 0.001 | [-0.023, 0.027] | 0.000 | 0.908 |
|  | Entorhinal | 0.051 | [0.010, 0.088] | 0.057 | 0.030 |
|  | Pars opercularis | 0.007 | [-0.015, 0.030] | 0.006 | 0.488 |
|  | Pars orbitalis | 0.009 | [-0.022, 0.038] | 0.006 | 0.472 |
|  | Pars triangularis | 0.010 | [-0.016, 0.035] | 0.007 | 0.445 |
|  | Rostral middle frontal | 0.009 | [-0.012, 0.027] | 0.011 | 0.315 |
|  | Superior frontal | 0.014 | [-0.003, 0.035] | 0.028 | 0.119 |
|  | Precentral gyrus | 0.026 | [0.003, 0.047] | 0.056 | 0.026 |
|  | Inferior temporal | 0.026 | [0.006, 0.048] | 0.088 | 0.012 |
|  | Middle temporal | 0.027 | [0.008, 0.039] | 0.098 | 0.008 |
|  | Superior temporal | 0.025 | [0.002, 0.043] | 0.057 | 0.027 |
|  | Transverse temporal | 0.030 | [-0.005, 0.058] | 0.031 | 0.060 |
|  | Insula | 0.039 | [0.017, 0.071] | 0.104 | 0.001* |
| Right | Caudal middle frontal | 0.020 | [0.002, 0.036] | 0.052 | 0.033 |
|  | Lateral orbitofrontal | 0.009 | [-0.021, 0.031] | 0.009 | 0.326 |
|  | Medial orbitofrontal | 0.025 | [0.005, 0.048] | 0.056 | 0.022 |
|  | Entorhinal | 0.063 | [0.000, 0.126] | 0.044 | 0.026 |
|  | Pars opercularis | 0.017 | [-0.004, 0.038] | 0.028 | 0.103 |
|  | Pars orbitalis | 0.010 | [-0.022, 0.032] | 0.008 | 0.528 |
|  | Pars triangularis | 0.010 | [-0.012, 0.025] | 0.009 | 0.331 |
|  | Rostral middle frontal | 0.006 | [-0.011, 0.020] | 0.008 | 0.418 |
|  | Superior frontal | 0.012 | [-0.008, 0.031] | 0.019 | 0.231 |
|  | Precentral gyrus | 0.028 | [0.005, 0.049] | 0.065 | 0.023 |
|  | Inferior temporal | 0.026 | [0.005, 0.051] | 0.072 | 0.011 |
|  | Middle temporal | 0.021 | [0.000, 0.038] | 0.061 | 0.052 |
|  | Superior temporal | 0.022 | [-0.033, 0.040] | 0.040 | 0.060 |
|  | Transverse temporal | 0.004 | [-0.034, 0.037] | 0.001 | 0.784 |
|  | Insula | 0.040 | [0.021, 0.063] | 0.167 | <0.001* |

*Note:* \* $p < 0.05$ , FDR-corrected.
BCa, bias-corrected and accelerated; CI, confidence interval; GLMM, general linear mixed model; NfL, neurofilament light chain protein; TRS, treatment-resistant schizophrenia.

Detailed results of GLMMs with main effects of plasma NfL separated by diagnostic group are available in Supplementary Tables S1 & S2.

## Discussion

This study investigated the utility of plasma NfL as a marker of neuroimaging abnormalities in a cohort of clozapine-treated TRS patients, compared to controls. The main finding is that plasma NfL levels predicted lower cortical thickness of the insula bilaterally among people with TRS, compared to healthy controls. Thus, the primary hypothesis regarding associations between plasma NfL and decreased thickness of discrete cortical areas was partially supported. These findings suggest that regional neurodegenerative changes may be occurring in a schizophrenia cohort that represents severe and chronic disease. Although the exact mechanism of how schizophrenia pathology may contribute to neuroimaging abnormalities is unclear, the association between NfL and neuroimaging abnormalities in the TRS group raises the possibility of a neurodegenerative course that specifically affects neuronal axons. To the best of our knowledge, this is the first study to explore plasma NfL in the context of neuroimaging in a large, well-characterised group of people with TRS (defined by current treatment with clozapine). We propose that plasma NfL, an established marker of neuronal injury and neurodegeneration, is associated with cortical thinning of key brain structures often implicated in schizophrenia.

The association of cortical thickness reductions of the right and left insula with plasma NfL in people with TRS versus controls was a key and novel finding. The insula is understood to have extensive connections to brain areas that process external sensory information, which if disrupted may contribute to a diminished ability to discriminate between internally- and externally-generated sensory information.(48) Misrepresentation of self-sources of speech as distinct and external has therefore been posited as a potential substrate for auditory hallucinations in schizophrenia.(49) Our findings are comparable with those of Van Erp et al.,(15) who demonstrated a discrete pattern of cortical thinning across the brain in schizophrenia compared to healthy controls, including the insula bilaterally. They further suggest that clozapine dose is associated with a higher degree of cortical reduction, while a reduced insular thickness corresponds to an earlier age of onset and greater duration of illness in people with schizophrenia. Volumetric reductions of the insula have even been observed in first-episode psychosis(50,51) as well as in individuals at ultra-high-risk of psychosis.(52) A functional MRI study also reported altered and poorly differentiated functional connectivity profiles between the anterior and posterior insulae and their targets in schizophrenia.(53) Diminished insula functional connectivity is also associated with longer illness duration which may suggest a progressive loss of connectivity of this network along the clinical course of the illness. Our finding of an association of plasma NfL levels and thinning of the insular cortex bilaterally may at least to some degree reflect aberrant connectivity of this region in schizophrenia.

Our results of a relationship between thinning of the insula bilaterally, and plasma NfL levels supports the notion of a neurodegenerative process occurring in severe forms of schizophrenia. Cortical thickness represents the density and arrangement of neurons, glial cells, dendritic processes, and synaptic spines.(54) Disruption to one or more of these neural components in localised areas of the brain may account for findings of cortical thinning associated with NfL among people with TRS compared to controls. Taken together, we propose the possibility of an active neurodegenerative process leading to axonal degeneration and injury, of which NfL is a marker, in patients who fail to respond adequately to antipsychotics. In addition to the conceptualisation of schizophrenia as a psychiatric illness, the current study adds weight to previous bodies of work speculating about the possibility of a neurodegenerative component.(6,55,56) These findings do not rule out the involvement of other neuropathological processes in severe schizophrenia, only that axonal injury may represent one mechanism driving these structural neuroimaging changes.

Our findings must be interpreted in the context of some limitations. Firstly, the cross-sectional design of the study meant that it was not possible to investigate whether longitudinal plasma NfL increases are associated with progressive brain changes in TRS. While progressive changes have been explored in schizophrenia previously,(5,16,17,57) no studies to date have examined such changes in association with NfL levels. Secondly, all participants with TRS recruited to the study were receiving clozapine and so it was not possible to disentangle pathological disease-driven brain changes from those arising due to, or accelerated by, clozapine use. Thirdly, the lack of specificity of NfL as a marker of neuronal insult and injury inherently limits the conclusions that can be drawn about the disease-driven pathogenic processes in schizophrenia. NfL is elevated in response to various neurodegenerative, inflammatory, vascular, and traumatic insults to neuronal axons.(24) Fourthly, the present study was unable to compare findings of the TRS group to a cohort of non-TRS patients in order to determine whether NfL-related neuroimaging abnormalities are a feature common across all schizophrenia patients, or whether they are unique to TRS. The utility of plasma NfL in predicting neuroimaging changes specific to patients exhibiting treatment-resistance was unexplored but should be a priority of future studies. Finally, participants recruited to the TRS cohort of our study were largely patients with long-standing illness. As a result, the present study was unable to investigate the clinical utility of NfL in detecting structural brain abnormalities in earlier stages of the schizophrenia disease course.

The current study provides evidence for an association between plasma NfL and reduced thickness in discrete brain structures, namely the bilateral insula, in a group of TRS patients compared to controls. These findings suggest that neuronal insult may be occurring in patients who respond inadequately to treatment, adding weight to a neurodegenerative model of schizophrenia and the notion of a subgroup of poor-outcome, severe schizophrenia represented by treatment-resistance. NfL and other markers of neurodegeneration are likely to be important correlates of clozapine-treated TRS patients. Large-scale prospective longitudinal studies are required to confirm findings presented in the current study. Future studies should also aim to appraise the clinical utility of plasma NfL in distinguishing TRS from non-TRS patients, as a marker of treatment response, as well as exploring NfL in other stages of the disorder and in at-risk cohorts.

## Data Availability

All data produced in the present study are available upon reasonable request to the authors

## Acknowledgements

The authors acknowledge the financial support of the CRC for Mental Health. The Cooperative Research Centre (CRC) programme is an Australian Government Initiative. The authors wish to acknowledge the CRC Scientific Advisory Committee, in addition to the contributions of study participants, clinicians at recruitment services, staff at the Murdoch Children’s Research Institute, staff at the Australian Imaging, Biomarkers and Lifestyle Flagship Study of Aging, and research staff at the Melbourne Neuropsychiatry Centre, including coordinators Merritt, A., Phassouliotis, C., and research assistants, Burnside, A., Cross, H., Gale, S., and Tahtalian, S.

Participants for this study were sourced, in part, through the Australian Schizophrenia Research Bank (ASRB), which is supported by the National Health and Medical Research Council of Australia (Enabling Grant N. 386500), the Pratt Foundation, Ramsay Health Care, the Viertel Charitable Foundation and the Schizophrenia Research Institute. We thank the Chief Investigators and ASRB Manager: Carr, V., Schall, U., Scott, R., Jablensky, A., Mowry, B., Michie, P., Catts, S., Henskens, F., Pantelis, C., Loughland, C. We acknowledge the help of Jason Bridge for ASRB database queries.

The authors are grateful for assistance from Brett Trounson and Dr Christopher Fowler and the team at The Florey Oak St Biobank. Finally, the authors would like to thank all the participants and their families. The corresponding author had full access to all the data in the study and had final responsibility for the decision to submit for publication.

## Author contributions

DE, OH, IE, CB, AFS, DV, CP developed conception and design of the work. DE, CW and OH carried out data collection, B-JC, DE, CW and CM performed data analysis. B-JC, DE, CW, CM, AFS, DV and CP made the interpretation of the results and B-JC wrote the first manuscript draft with assistance from DE, CW, CM, AFS and DV. SJ, OH, IE, CB, NT and CP provided critical revision of the manuscript. All authors approved the final version of the manuscript.

## References

1. Murray RM, Lewis SW. Is schizophrenia a neurodevelopmental disorder? Br Med J (Clin Res Ed). 1987;295(6600):681–682. doi:10.1136/bmj.295.6600.681

2. Jaaro-Peled H, Hayashi-Takagi A, Seshadri S, Kamiya A, Brandon NJ, Sawa A. Neurodevelopmental mechanisms of schizophrenia: understanding disturbed postnatal brain maturation through neuregulin-1-ErbB4 and DISC1. Trends Neurosci. 2009;32(9):485–495. doi:10.1016/j.tins.2009.05.007

3. Rapoport JL, Addington AM, Frangou S, Psych MRC. The neurodevelopmental model of schizophrenia: update 2005. Mol Psychiatry. 2005;10(5):434–449. doi:10.1038/sj.mp.4001642

4. Marenco S, Weinberger DR. The neurodevelopmental hypothesis of schizophrenia: following a trail of evidence from cradle to grave. Dev Psychopathol. 2000;12(3):501–527. doi:10.1017/s0954579400003138

5. Olabi B, Ellison-Wright I, McIntosh AM, Wood SJ, Bullmore E, Lawrie SM. Are there progressive brain changes in schizophrenia? A meta-analysis of structural magnetic resonance imaging studies. Biol Psychiatry. 2011;70(1):88–96. doi:10.1016/j.biopsych.2011.01.032

6. Pantelis C, Yücel M, Wood SJ, et al. Structural brain imaging evidence for multiple pathological processes at different stages of brain development in schizophrenia. Schizophr Bull. 2005;31(3):672–696. doi:10.1093/schbul/sbi034

7. Elkis H. Treatment-resistant schizophrenia. Psychiatr North Am. 2007;30(3):511–533. doi:10.1016/j.psc.2007.04.001

8. Suzuki T, Remington G, Mulsant BH, et al. Defining treatment-resistant schizophrenia and response to antipsychotics: a review and recommendation. Psychiatr Res. 2012;197(1-2):1–6. doi:10.1016/j.psychres.2012.02.013

9. Iasevoli F, Giordano S, Balletta R, et al. Treatment resistant schizophrenia is associated with the worst community functioning among severely-ill highly-disabling psychiatric conditions and is the most relevant predictor of poorer achievements in functional milestones. Prog Neuropsychopharmacol Biol Psychiatry. 2016;65:34–48. doi:10.1016/j.pnpbp.2015.08.010

10. Kahn RS, Sommer IE, Murray RM, et al. Schizophrenia. Nat Rev Dis Primer. 2015;1, 15067. doi:10.1038/nrdp.2015.67

11. Pantelis C, Velakoulis D, McGorry PD, et al. Neuroanatomical abnormalities before and after onset of psychosis: a cross-sectional and longitudinal MRI comparison. Lancet. 2003;361(9354):281–288. doi:10.1016/S0140-6736(03)12323-9

12. van Haren NE, Schnack HG, Cahn W et al. Changes in cortical thickness during the course of illness in schizophrenia. Arch Gen Psychiatry. 2011;68(9):871–880. doi:10.1001/archgenpsychiatry.2011.88

13. Vita A, Minelli A, Barlati S, et al. Treatment-Resistant Schizophrenia: Genetic and Neuroimaging Correlates. Front Pharmacol. 2019;10:402. doi:10.3389/fphar.2019.00402

14. Shepherd AM, Laurens KR, Matheson SL, Carr VJ, Green MJ. Systematic meta-review and quality assessment of the structural brain alterations in schizophrenia. Neurosci Biobehav Rev. 2012;36(4):1342–1356. doi:10.1016/j.neubiorev.2011.12.015

15. van Erp TGM, Walton E, Hibar DP, et al. Cortical Brain Abnormalities in 4474 Individuals With Schizophrenia and 5098 Control Subjects via the Enhancing Neuro Imaging Genetics Through Meta Analysis (ENIGMA) Consortium. Biol Psych. 2018;84(9):644–654. doi:10.1016/j.biopsych.2018.04.023

16. van Haren NE, Hulshoff Pol HE, Schnack HG, et al. Progressive brain volume loss in schizophrenia over the course of the illness: evidence of maturational abnormalities in early adulthood. Biol Psychiatry. 2008;63(1):106–113. doi:10.1016/j.biopsych.2007.01.004

17. Vita A, De Peri L, Deste G, Sacchetti E. Progressive loss of cortical gray matter in schizophrenia: a meta-analysis and meta-regression of longitudinal MRI studies. Transl Psychiatry. 2012;2(11):e190. doi:10.1038/tp.2012.116

18. Winkler AM, Kochunov P, Blangero J, et al. Cortical thickness or grey matter volume? The importance of selecting the phenotype for imaging genetics studies. Neuroimage. 2010;53(3):1135–1146. doi:10.1016/j.neuroimage.2009.12.028

19. Panizzon MS, Fennema-Notestine C, Eyler LT, et al. Distinct genetic influences on cortical surface area and cortical thickness. Cereb Cortex. 2009;19(11):2728–2735. doi:10.1093/cercor/bhp026

20. Weinberger DR. The neurodevelopmental origins of schizophrenia in the penumbra of genomic medicine. World Psychiatry. 2017;16(3):225–226. doi:10.1002/wps.20474

21. Cui Y, Liu B, Song M, et al. Auditory verbal hallucinations are related to cortical thinning in the left middle temporal gyrus of patients with schizophrenia. Psychol Med. 2018;48(1):115–122. doi:10.1017/S0033291717001520

22. Zhao Y, Zhang Q, Shah C, et al. Cortical Thickness Abnormalities at Different Stages of the Illness Course in Schizophrenia: A Systematic Review and Meta-analysis. JAMA Psychiatry. 2022;79(6):560–570. doi:10.1001/jamapsychiatry.2022.0799

23. Wannan CMJ, Cropley VL, Chakravarty MM, et al. Evidence for Network-Based Cortical Thickness Reductions in Schizophrenia. Am J Psychiatry. 2019;176(7):552–563. doi:10.1176/appi.ajp.2019.18040380

24. Khalil M, Teunissen CE, Otto M, et al. Neurofilaments as biomarkers in neurological disorders. Nat Rev Neurol. 2018;14(10):577–589. doi:10.1038/s41582-018-0058-z

25. Yuan A, Rao MV, Veeranna, Nixon RA. Neurofilaments and Neurofilament Proteins in Health and Disease. Cold Spring Harb Perspect Biol. 2017;9(4):a018309. Published 2017 Apr 3. doi:10.1101/cshperspect.a018309

26. Gaetani L, Blennow K, Calabresi P, Di Filippo M, Parnetti L, Zetterberg H. Neurofilament light chain as a biomarker in neurological disorders. J Neurol Neurosurg Psychiatry. 2019;90(8):870–881. doi:10.1136/jnnp-2018-320106

27. Ashton NJ, Janelidze S, Al Khleifat A, et al. A multicentre validation study of the diagnostic value of plasma neurofilament light. Nat Commun. 2021;12(1):3400. doi:10.1038/s41467-021-23620-z

28. Eratne D, Loi SM, Li QX, et al. Cerebrospinal fluid neurofilament light chain differentiates primary psychiatric disorders from rapidly progressive, Alzheimer’s disease and frontotemporal disorders in clinical settings. Alzheimers Dement. 2022;18(11):2218–2233. doi:10.1002/alz.12549

29. Eratne D, Kang M, Malpas C, et al. Plasma neurofilament light in behavioural variant frontotemporal dementia compared to mood and psychotic disorders. Aust N Z J Psychiatry. 2024;58(1):70–81. doi:10.1177/00048674231187312

30. Katisko K, Cajanus A, Jääskeläinen O, et al. Serum neurofilament light chain is a discriminative biomarker between frontotemporal lobar degeneration and primary psychiatric disorders. J Neurol. 2020;267(1):162–167. doi:10.1007/s00415-019-09567-8

31. Bavato F, Cathomas F, Klaus F, et al. Altered neuroaxonal integrity in schizophrenia and major depressive disorder assessed with neurofilament light chain in serum. J Psychiatr Res. 2021;140:141–148. doi:10.1016/j.jpsychires.2021.05.072

32. Rodrigues-Amorim D, Rivera-Baltanás T, Del Carmen Vallejo-Curto M, et al. Plasma β-III tubulin, neurofilament light chain and glial fibrillary acidic protein are associated with neurodegeneration and progression in schizophrenia. Sci Rep. 2020;10(1):14271. doi:10.1038/s41598-020-71060-4

33. Eratne D, Janelidze S, Malpas CB, et al. Plasma neurofilament light chain protein is not increased in treatment-resistant schizophrenia and first-degree relatives. Aust N Z J Psychiatry. 2022;56(10):1295–1305. doi:10.1177/00048674211058684

34. Mouchlianitis E, McCutcheon R, Howes OD. Brain-imaging studies of treatment-resistant schizophrenia: a systematic review. Lancet Psychiatry. 2016;3(5):451–463. doi:10.1016/S2215-0366(15)00540-4

35. Quarantelli M, Palladino O, Prinster A, et al. Patients with poor response to antipsychotics have a more severe pattern of frontal atrophy: a voxel-based morphometry study of treatment resistance in schizophrenia. Biomed Res Int. 2014;2014:325052. doi:10.1155/2014/325052

36. Anderson VM, Goldstein ME, Kydd RR, Russell BR. Extensive gray matter volume reduction in treatment-resistant schizophrenia. Int J Neuropsychopharmacol. 2015;18(7):pyv016. doi:10.1093/ijnp/pyv016

37. Sheehan DV, Lecrubier Y, Sheehan KH, et al. The Mini-International Neuropsychiatric Interview (M.I.N.I.): the development and validation of a structured diagnostic psychiatric interview for DSM-IV and ICD-10. J Clin Psychiatry. 1998;59 Suppl 20:22–57.

38. Fischl B, Salat DH, Busa E, et al. Whole brain segmentation: automated labeling of neuroanatomical structures in the human brain. Neuron. 2002;33(3):341–355. doi:10.1016/s0896-6273(02)00569-x

39. Fischl B, van der Kouwe A, Destrieux C, et al. Automatically parcellating the human cerebral cortex. Cereb Cortex. 2004;14(1):11–22. doi:10.1093/cercor/bhg087

40. Dale AM, Fischl B, Sereno MI. Cortical surface-based analysis. I. Segmentation and surface reconstruction. Neuroimage. 1999;9(2):179-194. doi:10.1006/nimg.1998.0395

41. Fischl B, Liu A, Dale AM. Automated manifold surgery: constructing geometrically accurate and topologically correct models of the human cerebral cortex. IEEE Trans Med Imaging. 2001;20(1):70–80. doi:10.1109/42.906426

42. Ségonne F, Pacheco J, Fischl B. Geometrically accurate topology-correction of cortical surfaces using nonseparating loops. IEEE Trans Med Imaging. 2007;26(4):518–529. doi:10.1109/TMI.2006.887364

43. Sled JG, Zijdenbos AP, Evans AC. A nonparametric method for automatic correction of intensity nonuniformity in MRI data. IEEE Trans Med Imaging. 1998;17(1):87–97. doi:10.1109/42.668698

44. Ségonne F, Dale AM, Busa E, et al. A hybrid approach to the skull stripping problem in MRI. Neuroimage. 2004;22(3):1060–1075. doi:10.1016/j.neuroimage.2004.03.032

45. Desikan RS, Ségonne F, Fischl B, et al. An automated labeling system for subdividing the human cerebral cortex on MRI scans into gyral based regions of interest. Neuroimage. 2006;31(3):968–980. doi:10.1016/j.neuroimage.2006.01.021

46. Manouchehrinia A, Piehl F, Hillert J, et al. Confounding effect of blood volume and body mass index on blood neurofilament light chain levels. Ann Clin Transl Neurol. 2020;7(1):139–143. doi:10.1002/acn3.50972

47. Benjamin Y, Hochberg Y. Controlling the false discovery rate: a practical and powerful approach to Multiple testing. J R Stat Soc. 1995;57:289–300

48. Wylie KP, Tregellas JR. The role of the insula in schizophrenia. Schizophr Res. 2010;123(2-3):93–104. doi:10.1016/j.schres.2010.08.027

49. Palaniyappan L, Balain V, Radua J, Liddle PF. Structural correlates of auditory hallucinations in schizophrenia: a meta-analysis. Schizophr Res. 2012;137(1-3):169–173. doi:10.1016/j.schres.2012.01.038

50. Takahashi T, Wood SJ, Soulsby B, et al. Follow-up MRI study of the insular cortex in first-episode psychosis and chronic schizophrenia. Schizophr Res. 2009;108(1-3):49–56. doi:10.1016/j.schres.2008.12.029

51. Takahashi T, Wood SJ, Soulsby B, et al. Diagnostic specificity of the insular cortex abnormalities in first-episode psychotic disorders. Prog Neuropsychopharmacol Biol Psychiatry. 2009;33(4):651–657. doi:10.1016/j.pnpbp.2009.03.005

52. Takahashi T, Wood SJ, Yung AR, et al. Insular cortex gray matter changes in individuals at ultra-high-risk of developing psychosis. Schizophr Res. 2009;111(1-3):94–102. doi:10.1016/j.schres.2009.03.024

53. Tian Y, Zalesky A, Bousman C, Everall I, Pantelis C. Insula Functional Connectivity in Schizophrenia: Subregions, Gradients, and Symptoms. Biol Psychiatry Cogn Neurosci Neuroimaging. 2019;4(4):399–408. doi:10.1016/j.bpsc.2018.12.003

54. Selemon LD, Goldman-Rakic PS. The reduced neuropil hypothesis: a circuit based model of schizophrenia. Biol Psychiatry. 1999;45(1):17–25. doi:10.1016/s0006-3223(98)00281-9

55. Kochunov P, Hong LE. Neurodevelopmental and neurodegenerative models of schizophrenia: white matter at the center stage. Schizophr Bull. 2014;40(4):721–728. doi:10.1093/schbul/sbu070

56. Lieberman JA. Is schizophrenia a neurodegenerative disorder? A clinical and neurobiological perspective. Biol Psychiatry. 1999;46(6):729–739. doi:10.1016/s0006-3223(99)00147-x

57. Andreasen NC, Nopoulos P, Magnotta V, Pierson R, Ziebell S, Ho BC. Progressive brain change in schizophrenia: a prospective longitudinal study of first-episode schizophrenia. Biol Psychiatry. 2011;70(7):672–679. doi:10.1016/j.biopsych.2011.05.017

