## Supplementary Table S1 for "Associations between structural brain changes and blood neurofilament light chain protein in treatment-resistant schizophrenia"

**Supplementary Table S1.** General linear mixed models for control participants with corresponding main effects of plasma NfL for each brain structure thickness

| **Hemisphere** | **GLMM** | **Main effects of plasma NfL** | | | |
| --- | --- | --- | --- | --- | --- |
|  |  | B* | BCa 95% CI | *η^2^_p_* | *p*-value |
| Left | Caudal middle frontal | 0.010 | [−0.007, 0.27] | 0.034 | 0.247 |
|  | Lateral orbitofrontal | 0.008 | [−0.007, 0.023] | 0.029 | 0.289 |
|  | Medial orbitofrontal | −0.004 | [−0.021, 0.013] | 0.006 | 0.630 |
|  | Entorhinal | 0.042 | [0.008, 0.076] | 0.136 | 0.018 |
|  | Pars opercularis | 0.011 | [−0.008, −0.002] | 0.210 | 0.003 |
|  | Pars orbitalis | 0.011 | [−0.013, 0.034] | 0.021 | 0.361 |
|  | Pars triangularis | 0.017 | [−0.004, 0.037] | 0.066 | 0.106 |
|  | Rostral middle frontal | 0.016 | [0.001, 0.031] | 0.109 | 0.006 |
|  | Superior frontal | 0.016 | [0.001, 0.031] | 0.102 | 0.042 |
|  | Precentral gyrus | 0.029 | [0.011, 0.046] | 0.216 | **0.002** |
|  | Inferior temporal | 0.019 | [0.001, 0.036] | 0.109 | 0.035 |
|  | Middle temporal | 0.021 | [0.007, 0.036] | 0.191 | **0.004** |
|  | Superior temporal | 0.027 | [0.011, 0.043] | 0.228 | **0.002** |
|  | Transverse temporal | 0.034 | [0.005, 0.062] | 0.125 | 0.023 |
|  | Insula | 0.023 | [0.003, 0.042] | 0.125 | 0.023 |
| Right | Caudal middle frontal | 0.013 | [−0.001, 0.027] | 0.085 | 0.064 |
|  | Lateral orbitofrontal | 0.001 | [−0.017, 0.018] | 0.000 | 0.940 |
|  | Medial orbitofrontal | 0.013 | [−0.004, 0.029] | 0.058 | 0.130 |
|  | Entorhinal | 0.025 | [−0.025, 0.074] | 0.026 | 0.318 |
|  | Pars opercularis | 0.019 | [0.001, 0.038] | 0.103 | 0.040 |
|  | Pars orbitalis | 0.001 | [−0.019, 0.022] | 0.000 | 0.890 |
|  | Pars triangularis | 0.017 | [−0.001, 0.036] | 0.086 | 0.062 |
|  | Rostral middle frontal | 0.013 | [0.001, 0.025] | 0.112 | 0.033 |
|  | Superior frontal | 0.010 | [−0.006, 0.026] | 0.042 | 0.197 |
|  | Precentral gyrus | 0.030 | [0.012, 0.047] | 0.235 | **0.001** |
|  | Inferior temporal | 0.012 | [−0.005, 0.028] | 0.052 | 0.153 |
|  | Middle temporal | 0.017 | [0.002, 0.031] | 0.122 | 0.025 |
|  | Superior temporal | 0.022 | [0.004, 0.039] | 0.140 | 0.016 |
|  | Transverse temporal | 0.009 | [−0.022, 0.039] | 0.009 | 0.563 |
|  | Insula | 0.020 | [0.005, 0.036] | 0.150 | 0.012 |

*Note:* **Bolded values are significant at *p* < 0.05, FDR-corrected.** CI, confidence interval; GLMM, general linear mixed model; NfL, neurofilament light chain protein; TRS, treatment-resistant schizophrenia.
