## Supplementary Table S2 for "Associations between structural brain changes and blood neurofilament light chain protein in treatment-resistant schizophrenia"

**Supplementary Table S2.** General linear mixed models for TRS participants with corresponding main effects of plasma NfL for each brain structure thickness.

| **Hemisphere** | **GLMM** | **Main effects of plasma NfL** | | | |
| --- | --- | --- | --- | --- | --- |
|  |  | B* | 95% CI | *η^2^_p_* | *p*-value |
| Left | Caudal middle frontal | −0.006 | [−0.028, 0.015] | 0.010 | 0.553 |
|  | Lateral orbitofrontal | −0.006 | [−0.026, 0.013] | 0.013 | 0.502 |
|  | Medial orbitofrontal | 0.005 | [−0.015, 0.024] | 0.006 | 0.641 |
|  | Entorhinal | −0.034 | [−0.084, 0.016] | 0.051 | 0.178 |
|  | Pars opercularis | −0.009 | [−0.031, 0.014] | 0.018 | 0.432 |
|  | Pars orbitalis | 0.002 | [−0.026, 0.031] | 0.001 | 0.863 |
|  | Pars triangularis | −0.007 | [−0.033, 0.020] | 0.008 | 0.605 |
|  | Rostral middle frontal | −0.003 | [−0.023, 0.016] | 0.003 | 0.729 |
|  | Superior frontal | −0.001 | [−0.019, 0.018] | 0.000 | 0.932 |
|  | Precentral gyrus | −0.016 | [−0.041, 0.009] | 0.045 | 0.208 |
|  | Inferior temporal | −0.013 | [−0.030, 0.004] | 0.068 | 0.118 |
|  | Middle temporal | −0.015 | [−0.033, 0.003] | 0.076 | 0.099 |
|  | Superior temporal | −0.012 | [−0.037, 0.012] | 0.029 | 0.310 |
|  | Transverse temporal | −0.008 | [−0.046, 0.030] | 0.005 | 0.670 |
| Right | Insula | −0.027 | [−0.053, −0.001] | 0.109 | 0.046 |
|  | Caudal middle frontal | −0.014 | [−0.034, 0.006] | 0.054 | 0.168 |
|  | Lateral orbitofrontal | −0.013 | [−0.035, 0.009] | 0.041 | 0.232 |
|  | Medial orbitofrontal | −0.020 | [−0.045, 0.005] | 0.071 | 0.111 |
|  | Entorhinal | −0.055 | [−0.117, 0.006] | 0.087 | 0.076 |
|  | Pars opercularis | −0.007 | [−0.028, 0.015] | 0.011 | 0.539 |
|  | Pars orbitalis | 0.006 | [−0.019, 0.030] | 0.006 | 0.642 |
|  | Pars triangularis | 0.006 | [−0.019, 0.031] | 0.007 | 0.626 |
|  | Rostral middle frontal | 0.004 | [−0.013, 0.021] | 0.007 | 0.633 |
|  | Superior frontal | −0.006 | [−0.024, 0.013] | 0.012 | 0.524 |
|  | Precentral gyrus | −0.011 | [−0.037, 0.015] | 0.021 | 0.395 |
|  | Inferior temporal | −0.020 | [−0.042, 0.003] | 0.084 | 0.082 |
|  | Middle temporal | −0.008 | [−0.026, 0.010] | 0.022 | 0.379 |
|  | Superior temporal | −0.004 | [−0.031, 0.022] | 0.003 | 0.729 |
|  | Transverse temporal | −0.006 | [−0.046, 0.033] | 0.003 | 0.745 |
|  | Insula | −0.024 | [−0.044, −0.004] | 0.146 | 0.019 |

*Note:* **Bolded values are significant at *p* < 0.05, FDR-corrected.** CI, confidence interval; GLMM, general linear mixed model; NfL, neurofilament light chain protein; TRS, treatment-resistant schizophrenia.
