## Supplementary Figure S2 for "Associations between structural brain changes and blood neurofilament light chain protein in treatment-resistant schizophrenia"

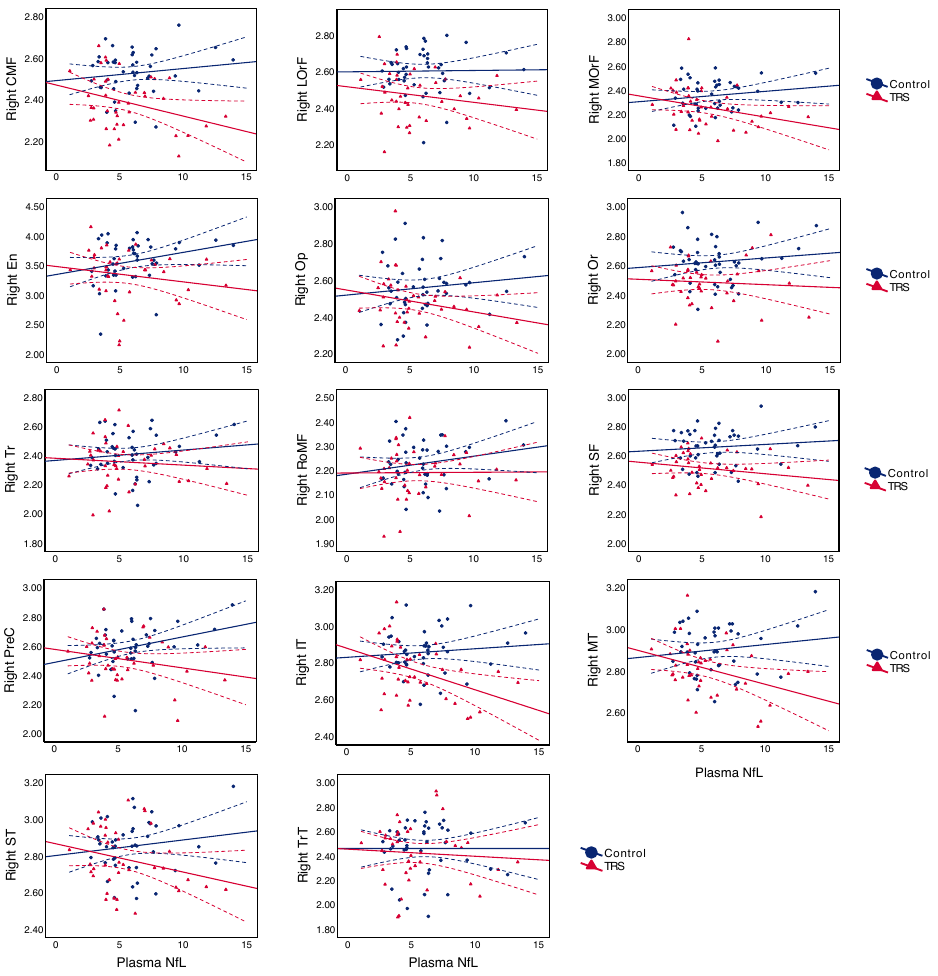

**Supplementary Figure S2.** Interaction plots with trendlines and 95% confidence intervals for non-significant right hemisphere brain thicknesses of interest. *Note:* Plasma NfL in pg/mL; thickness in mm. CMF, caudal middle frontal; En, entorhinal; IT, inferior temporal; LOrF, lateral orbitofrontal; MOrF, medial orbitofrontal; MT, middle temporal; Op, pars opercularis; Or, pars orbitalis; PreC, precentral gyrus; RoMF, rostral middle frontal; SF, superior frontal; ST, superior temporal; Tr, pars triangularis; TrT, transverse temporal.
